# Efficient high throughput SARS-CoV-2 testing to detect asymptomatic carriers

**DOI:** 10.1101/2020.04.14.20064618

**Authors:** Noam Shental, Shlomia Levy, Vered Wuvshet, Shosh Skorniakov, Yonat Shemer-Avni, Angel Porgador, Tomer Hertz

## Abstract

The COVID-19 pandemic is rapidly spreading throughout the world. Recent reports suggest that 10-30% of SARS-CoV-2 infected patients are asymptomatic. Other studies report that some subjects have significant viral shedding prior to symptom onset. Since both asymptomatic and pre-symptomatic subjects can spread the disease, identifying such individuals is critical for effective control of the SARS-CoV-2 pandemic. Therefore, there is an urgent need to increase diagnostic testing capabilities in order to also screen asymptomatic carriers. In fact, such tests will be routinely required until a vaccine is developed. Yet, a major bottleneck of managing the COVID-19 pandemic in many countries is diagnostic testing, due to limited laboratory capabilities as well as limited access to genome-extraction and Polymerase Chain Reaction (PCR) reagents. We developed P-BEST - a method for Pooling-Based Efficient SARS-CoV-2 Testing, using a non-adaptive group-testing approach, which significantly reduces the number of tests required to identify all positive subjects within a large set of samples. Instead of testing each sample separately, samples are pooled into groups and each pool is tested for SARS-CoV-2 using the standard clinically approved PCR-based diagnostic assay. Each sample is part of multiple pools, using a combinatorial pooling strategy based on compressed sensing designed for maximizing the ability to identify all positive individuals. We evaluated P-BEST using leftover samples that were previously clinically tested for COVID-19. In our current proof-of-concept study we pooled 384 patient samples into 48 pools providing an 8-fold increase in testing efficiency. Five sets of 384 samples, containing 1-5 positive carriers were screened and all positive carriers in each set were correctly identified. P-BEST provides an efficient and easy-to-implement solution for increasing testing capacity that will work with any clinically approved genome-extraction and PCR-based diagnostic methodologies.

## Introduction

The COVID-19 pandemic is rapidly spreading throughout the world. Recent reports suggest that 10-30% of SARS-CoV-2 infected patients are asymptomatic^1-3^. Other studies report that some subjects have significant viral shedding prior to symptom onset^4^. Since both asymptomatic and pre-symptomatic subjects can spread the disease^1,2^, identifying such individuals is critical for effective control of the SARS-CoV-2 pandemic. A major bottleneck of managing the COVID-19 pandemic in many countries is diagnostic testing which is primarily performed on symptomatic patients, due to limited laboratory capabilities as well as limited access to genome-extraction and Polymerase Chain Reaction (PCR) reagents. Hence, there is an urgent need to increase diagnostic testing capabilities in order to allow screening of asymptomatic populations which contribute to disease spread. In fact, such tests will be routinely required until a vaccine is developed. If the percentage of carriers in the tested set of samples is sufficiently low (∼1%), the method can correctly identify all positive individuals using a much smaller number of diagnostic tests as compared to testing each individual sample separately.

## Methods

We developed P-BEST - a method for <u>P</u>ooling-<u>B</u>ased <u>E</u>fficient <u>S</u>ARS-CoV-2 <u>T</u>esting, using a group-testing approach, which significantly reduces the number of tests required to identify all positive subjects within a large set of samples (**Figure 1**). Instead of testing each sample separately, samples are pooled into groups and each pool is tested for SARS-CoV-2 using the standard clinically approved PCR-based diagnostic assay. Each sample is part of multiple pools, using a combinatorial pooling strategy designed for maximizing the ability to identify all positive individuals (**Figure 1**) ^5,6^. If the percentage of carriers in the tested set of samples is sufficiently low (∼1%), the method can correctly identify all positive individuals using a much smaller number of diagnostic tests as compared to testing each individual sample separately. In our current proof-of-concept study of P-BEST, we pooled 384 patient samples into 48 pools, each containing 48 samples. Each sample was added to six different pools. Pools were designed based on a Reed-Solomon error correcting code, and were generated using an automated liquid dispensing robot. Following genome-extraction and PCR amplification of all pools, a decoding algorithm was used to identify carriers that were then tested individually for verification. The total time for pooling 384 samples into 48 pools using a basic liquid dispensing robot (Arise EZMate-601) was <5 hours, and was performed in a standard BSL-2 laboratory, since individual samples were diluted in a lysis buffer that inactivates all viral particles.

**Figure 1.**
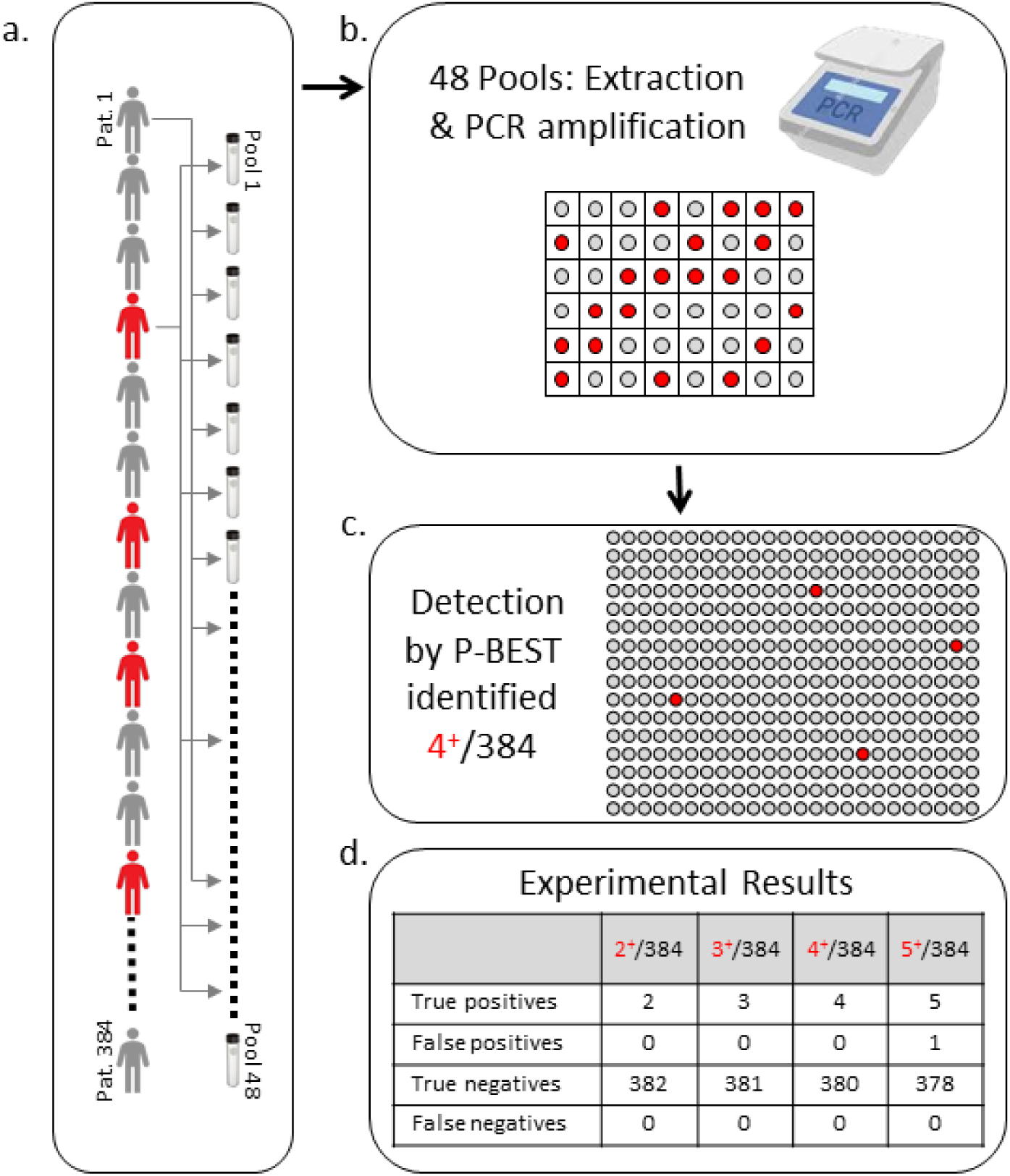
P-BEST design and detection results obtained for a set of 384 samples that contains 4 infected subjects. **a**. Generating 48 pools from 384 samples based on P-BEST; each sample is distributed to six pools. Subjects in red represent the four unidentified infected individuals within the set of 384 samples. Pools are generated using a combinatorial pooling design based on an error correcting code, that optimizes carrier detection. **b**. Extracting genetic material from the pools followed by standard PCR amplification. Positive pools are designated by red circles. **c**. P-BEST identifies the positive samples out of the 384 samples using an optimization-based algorithm. **d**. Results of the four experiments performed, containing 2, 3, 4 or 5 positive samples, respectively. No errors occurred for two to four positive samples, while for five positives a single false positive sample was added to the true positive ones. Details of the pooling design and the decoding method are provided in the supplementary online materials.

## Results

We evaluated P-BEST using leftover samples that were previously clinically tested for COVID-19. Samples diluted in lysis buffer were pooled into 48 pools, each containing a set of 48 unique samples. Pooled samples were then tested by the clinical diagnostic laboratory of the Soroka University Medical Center using a clinically approved COVID-19 PCR-based diagnostic protocol that included an RNA extraction stage. We tested P-BEST using four sets of 384 samples, each containing an increasing number of positive carriers ranging from two to five. We found that P-BEST was able to correctly identify all positive carriers within these four sets of 384 samples using only 48 tests per set, providing an 8-fold increase in testing efficiency. Simulations demonstrated that the method can correctly identify up to 5/384 (1.3%) of carriers, with an average number of false positives that was less than 2.75, and an average number of false negatives that was less than 0.33.

## Discussion

EST provides an efficient and easy-to-implement solution for increasing testing capacity that will work with any clinically approved genome-extraction and PCR-based diagnostic methodologies. Its implementation only requires the use of a widely available automated liquid dispensing robot. Importantly, our results demonstrate that P-BEST can use non-infectious samples (diluted in lysis buffer), allowing the automated pooling to be performed in BSL-2 laboratories. While our current pooling design provides an 8-fold increase in testing efficiency, this can be further improved by increasing the number of samples per pool. Preliminary experiments have demonstrated that a positive sample can be detected even within a pool of 128 subjects, which would allow efficient testing of >1000 samples. P-BEST is optimized for efficiently testing populations with low carrier frequencies (1.3% in this case). It is therefore best suited for screening asymptomatic populations that are not at high risk of being infected, excluding for example subjects who were in close contact with confirmed SARS-CoV-2 cases. Importantly, if the carrier frequency is above 1.3%, P-BEST will lose efficiency, but in turn may help identifying SARS-CoV-2 hotspots. Code and protocols required for implementation of P-BEST can be found at https://github.com/NoamShental/PBEST.

## Data Availability

all code is publicly available on github

https://github.com/NoamShental/PBEST

## Acknowledgements

We thank Avishai Edri, Aner Ottolenghi and Yariv Greenshpan for helping with sample preparation and RNA measurements. We thank Dr. Rachel Steinberg for help with diagnostic testing. We thank Jenny Racah and Shelly Levy-Tzedek for constructive discussions and manuscript editing. Noam Shental would like to thank Dalia Shental and Menashe Sobel for useful discussions. Noam Shental, Tomer Hertz and Angel Pogrador equally contributed to this manuscript.

## Supplementary Materials

### Methods

#### SARS Cov-2 clinical diagnostics

Naso and oropharynx (NP) swabs were collected from patients with symptoms of respiratory infection for analysis by the laboratory of Clinical Virology in Soroka University Medical Center (SUMC), which is approved by the Israeli Ministry of Health to test for SARS-CoV-2 infections. The laboratory employs the clinically approved SARS-CoV-2 detection kits of SeeGene (California, USA) for both viral nucleic acid extraction and qRT-PCR-based amplification for 3 SARS-CoV-19 genes (STARMag and 2019-nCoV Assay kits, respectively, done in the STARlet robot). Samples were analyzed individually, and positive and negative results were recorded. To neutralize the viral infectivity, 500µl of the sample is combined with 350µl of lysis buffer of the STARMag kit; following 20’of incubation, 350µl were used for nucleic acid extraction into a volume of 100µl extracted genome, from which 8µl were taken for the 2019-nCoV PCR assay.

#### Clinical samples for P-BEST

Frozen leftover media-lysis buffer (near 500µl) from clinically tested samples were thawed and then used to re-test the samples using the P-BEST approach. To test the sensitivity of the method, we generated several sets of 384 samples each containing 2-5 positive samples (Set1-4, respectively). Pools were prepared using a liquid handling robot (Arise EZMATE-601) using a code written in Python. The code automatically generates a command file for the robot to use. Samples were manually pipetted into 96 well plates from which the robot assembled a set of 48 pools, each containing 48 distinct samples. We generated 48 pools for each sample set. Each pool contained equal volumes from 48 samples (11µl /sample, 528µl per pool). Each individual sample was represented in six different pools. Analogously to single samples, 350µl from each pool were then used for nucleic acid extraction into a volume of 100µl, using the clinically approved STARMag kit and the STARlet robot. Then, 8µl were used for qRT-PCR to detect the E gene of SARS-CoV-2, based on a method that was clinically validated and employed in the SUMC-clinical virology laboratory prior to the introduction of the 2019-nCoV SeeGene kit. We used this PCR method due to shortage in the 2019-nCoV Assay kits of SeeGene; yet, to verify competence with the newly approved 2019-nCoV Assay kit, we re-validated positive pools with the See-Gene kit.

#### PCR sensitivity of pooled samples

Since in P-Best samples are diluted into pools of 48 subjects, there is an inherent drop in PCR sensitivity of about 5-6 cycles (a factor of 2^5^-2^6^), which was indeed observed experimentally and may result in false negative pools. In retrospective analysis, we found that only a single pool that included a carrier yielded a negative PCR result (out of expected 70 positive pools across four experiments). Similarly, only 1/122 of the negative pools was positively identified by the PCR-based assay. These two errors had no effect on the detection capabilities of P-BEST, which is robust to both of these types of errors.

Since our proof-of-concept experiments used samples from positive individuals that were previously identified, we observed an additional 5-cycle drop in PCR sensitivity that was likely caused by RNA degradation of samples following freeze-thaw cycles. To confirm this, we tested several individual samples before and after a freeze-thaw cycle and observed a similar drop in PCR sensitivity. However, P-BEST was still able to correctly identify all positive subjects within each of the sample sets. We note that this reduction will not occur when P-BEST will be implemented on fresh samples collected directly into lysis buffer, per our current experimental protocol. To further address the sensitivity issues we propose to (1) modify the sample collection protocol to collect samples directly into lysis buffer, thereby increasing the initial concentration of RNA in the collected samples; and (2) Test P-BEST pooled samples using the SeeGene SARS-CoV-2 PCR kit, that also includes primers for the NP SARS-COV-2 gene, which in our preliminary experiments had higher sensitivity as compared to the other genes included in the kit, and was positive even when diluting a single positive sample into a pool of 120 subjects (Supplemental Figure 1A).

**Supplemental Figure 1:**
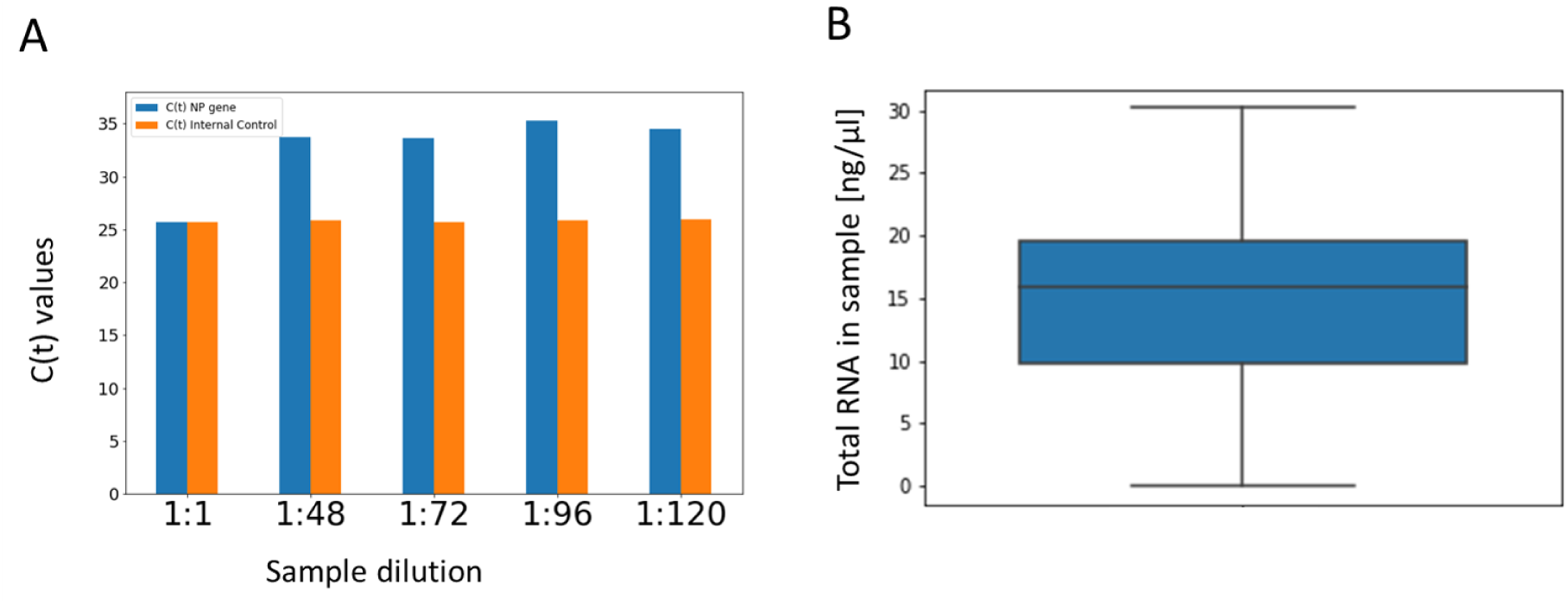
Evaluating pooling sensitivity and variation in total RNA levels. (A) A positive sample was serially diluted into a pool of 48 negative subjects, simulating the effect of RNA dilution of the sample in pools of increasing size ranging from 48 to 120. RNA was extracted from each diluted pooled sample, followed by PCR detection using the See-Gene clinical diagnostic SARS-CoV-2 kit. The figure shows the C(t) values of the NP gene for each dilution. (B) A boxplot of the total RNA levels of 48 single samples, as measured by the Qubit RNA IQ assay (Invitrogen). The line denotes the median value and box limits are the 25th and 75th percentiles. Lines denote the 95% confidence intervals.

#### Potential applications of P-BEST

P-BEST is best suited for screening asymptomatic populations, in which the carrier frequency may be sufficiently low to allow efficient detection. Such scenarios include routine screens of health-care workers and staff in nursing homes, as well population screens in different regions to identify new hotspots of the SARS-CoV-2 spread as early as possible. We note that additional pooling designs may be implemented to estimate the carrier frequency of SARS-CoV-2 in a given population, that may identify the specific scenarios in which P-BEST will be useful.

#### Group testing and sample pooling method

The mathematical field of group testing^7^ aims to tackle the problem of identifying individuals carrying a certain rare trait out of a large population by designing an efficient set of pools and measuring each pool as if it were a single sample. In general, pooling is designed in such a way that each individual has a unique ‘footprint’ on the set of pools, thus allowing carrier identification. Group testing dates back to the mid-20th century^8^ and since then many intricate pooling designs have been described and rigorously analyzed. Group testing has been successfully applied in data compression^9^, computation in the data stream model^10^ and in molecular biology^11,12^. In our previous studies we described a combination of group testing and next generation sequencing for detecting carriers of rare genetic mutations. We detected all individual carriers of rare mutations out of a set of 1024 mutagenized *Sorghum bicolor* plants using a set of 48 pools^5,6^.

The efficiency of group testing, often measured by the ratio between the number of screened individuals to the number of pools, generally increases with the decreasing frequency of the observed trait. For example, screening for a trait that appears in 0.1% of the population can be done more efficiently than screening for a trait that appears in 1% of the population. Moreover, when the carrier rate exceeds ∼5% group testing is no longer effective, since the required number of pools would be comparable to the number of samples tested.

To optimize efficiency, the pooling design needs to be tailored to the expected carrier rate. If the true carrier rate exceeds the expected rate, the method will identify larger sets of suspected carriers, which may include false positives, and in some cases may also fail to identify some of the carriers (false negatives). Therefore it is imperative to evaluate the robustness of a specific pooling design to higher carrier rates (e.g., Figure 2, and Supplemental Figures 2-3).

**Figure 2.**
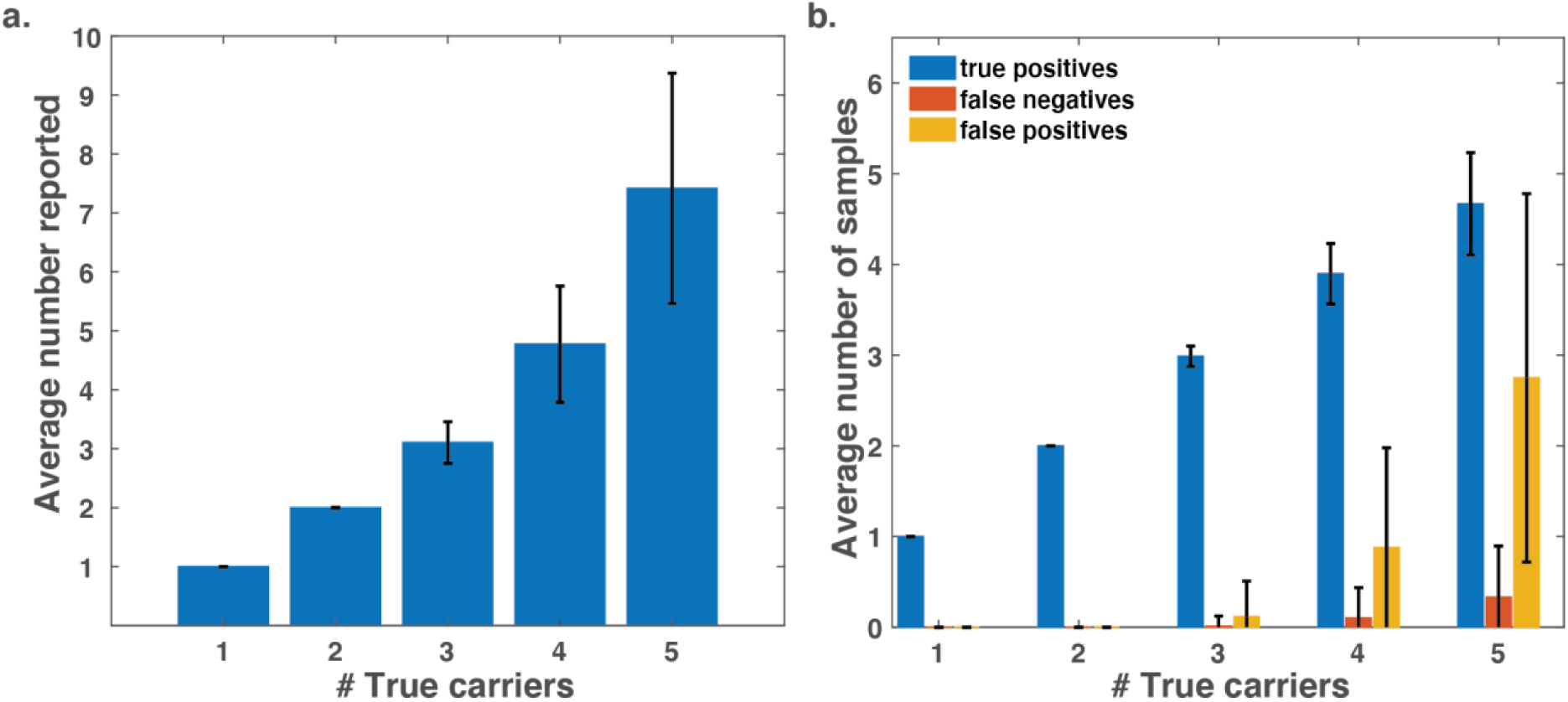
P-BEST in silico performance. A small number of carriers were randomly assigned to 384 samples and a P-BEST experiment was simulated using the reported pooling design. Simulations accounted for the estimated variation in RNA amounts based on measurements of n=48 individual samples, and also assumed that one of the 48 pools failed PCR amplification. Samples reported by P-BEST were compared to the true simulated sample labels to estimate P-BEST’s success rate. Results were averaged over 3000 simulations. (**a**) The average number of samples reported by P-BEST as a function of the number of true carriers. For example, P-BEST reports exactly two samples when simulating two carriers, and retrieves an average of ∼7.4 samples when the simulated set containing five carriers. (**b**) The average number of true positives, false negatives and false positives identified for a given number of simulated carriers. Even for five the number of false negatives is lower than one, and the average number of false positives remains low (<3), i.e., allowing only a minor increase in the number of required tests.

Our current detection strategy is based on binary results from each pool, i.e., a pool is either amplified or not. Since PCR results are quantitative, it may be possible to use this information in the P-BEST reconstruction algorithm to also estimate the C(t) values of each carrier (which is proportional to the individual’s viral load and may be clinically relevant).

#### P-BEST detection algorithm

Carrier detection was performed using the Gradient Projection for Sparse Reconstruction (GPSR) algorithm^13^ as in our former studies^5,6^. The transformation from fractional to discrete results was done using the following algorithm: The 20 samples with highest scores were selected and only subsets of these 20 were further considered. In total *2*^*20*^ subsets of samples were tested. Each subset corresponds to a vector *x* of length 384, in which the entries of the selected samples were equal to 1, and all others were set to zero. The product of *Mx*, where *M* is the pooling matrix, was compared to the measurement vector *y*. The vector for which ‖*Mx* − *y*‖ achieved its minimum was selected.

#### P-BEST robustness

To test the robustness of P-BEST we considered two types of potential noise factors. First, variation in initial RNA levels may cause samples to ‘disappear’ from all or part of the pools. Variation in RNA levels were estimated from Qubit measurements of 48 samples. The average RNA concentration was 15ng/µl, with standard deviation of 7ng/µl (Supplemental Figure 1B). These values were used in our simulations. A second possible source of noise is due to PCR amplification, which may fail in a certain number of pools. Supplemental Figures 2-3 display the effects of these two noise factors on our decoding accuracy. The average number of falsely detected samples – both false negatives and false positives does not change significantly. Hence the total number of required experiments remains unchanged even in highly noisy scenarios.

**Supplemental Figure 2:**
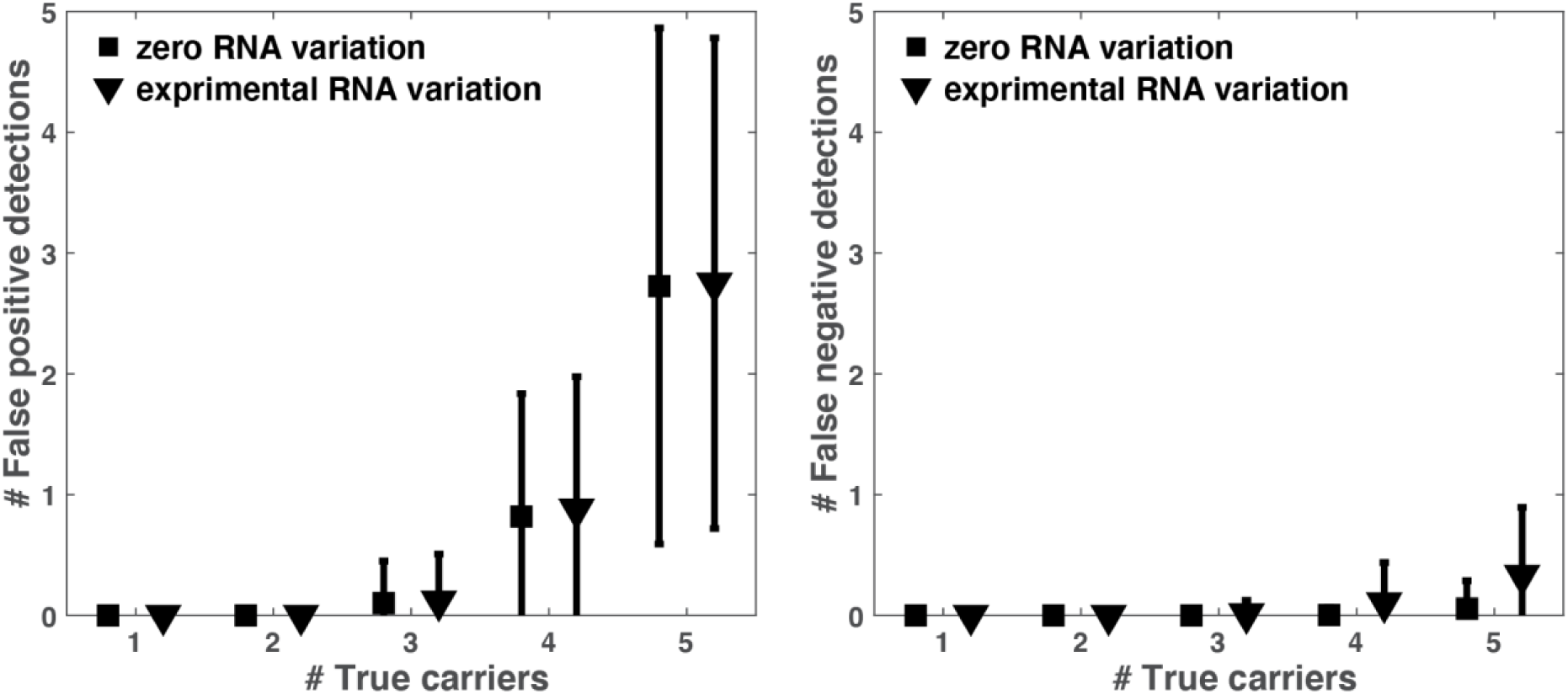
Evaluating the effect of variation in RNA levels on P-BEST performance. To assess the effects of variations in RNA levels, we measured the average number of false positive and false negative detections as a function of the true number of carriers across 3000 simulations in two scenarios: (1) No noise in RNA levels (black square) and (2) RNA noise based on the measured variation of RNA levels across 48 samples (see Supplemental Figure 1B). The false positive (left panel) and false negative (right panel) detection rates for the two scenarios show that RNA variation does not significantly degrade P-BEST performance. All simulations considered one dropped pool.

**Supplemental Figure 3:**
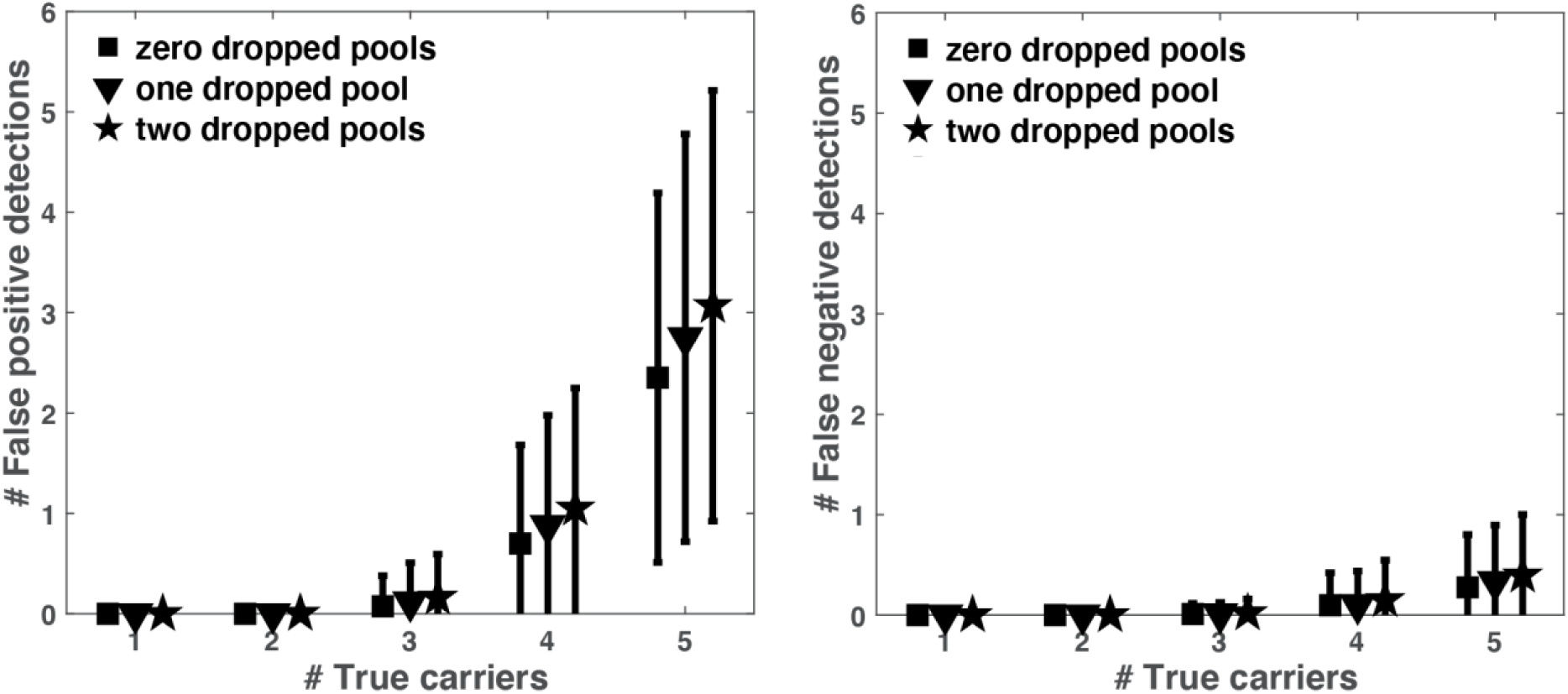
Evaluating the effect of dropped pools on P-BEST performance. To assess the effects of dropped pools due to PCR amplification failures, we measured the average number of false positive (left panel) and false negative (right panel) detections as a function of the true number of carriers across 3000 simulations for zero, one or two randomly dropped pools. P-BEST seems to be robust to 1-2 dropped pools. All simulations considered the experimental level of RNA variation as measured across 48 samples (Supplemental Figure 1B).

#### SARS-CoV-2 asymptomatic carriers and their role in disease transmission

There have been multiple recent studies that identified asymptomatic carriers and have attempted to estimate their rate and contribution to disease spread. A study of 2685 tourists in the New York area conducted over two seasons found that 6.2% of subjects tested positive for at least one respiratory virus, and 38.7% of these were infected with circulating Human Corona viruses^14^. Rothe et al. described transmission of SARS-CoV-2 from a German patient who was infected by a Chinese businesswoman who visited Germany^15^. Importantly, she was asymptomatic during her visit to Germany, and only developed symptoms after returning to China. Two other German co-workers were infected but only came in contact with the German patient who was asymptomatic. Mizumoto et al.^3^ reported that 50.5% of infected patients on board the Diamond Princess cruise ship were asymptomatic at time of diagnosis. Using a model, they estimated that the estimated asymptomatic proportion (among all infected cases) was 17.9% (95%CrI: 15.5–20.2%). Another study of evacuated Japanese nationals from Wuhan China, estimated that 30.8% of subjects were asymptomatic (95% CI: 7.7%-53.8%)^16^.

